# Diagnostic Accuracy of the Daye Diagnostic Tampon Compared to Clinician-Collected and Self-Collected Vaginal Swabs for Detecting HPV: A Comparative Study

**DOI:** 10.1101/2024.12.02.24318200

**Authors:** Valentina Milanova, Michelle Gomes, Kalina Mihaylova, John Luke Twelves, Jan Multmeier, Hana McMahon, Hannah McCulloch, Kate Cuschieri

## Abstract

**Introduction:** Cervical cancer screening is vital for achieving global elimination of this preventable disease. Vaginal self-sampling (VSS) for human papillomavirus (HPV) has the potential to increase screening uptake, particularly among individuals who may be underserved by clinician collection. Expanding self-sampling options with accurate, acceptable collection devices is essential. The Daye Diagnostic Tampon (DDT) offers an innovative approach, utilising a menstrual tampon to collect vaginal and cervical fluid for HPV screening. This study assessed the diagnostic accuracy of the DDT in detecting high-risk HPV infections, using clinician-collected swabs (CCS) as the reference standard.

**Methods:** In this UK-based study, 260 participants provided CCS, VSS, and DDT samples for HPV testing. Samples were analysed using a clinically validated assay for 14 high-risk HPV types. Sensitivity, specificity, positive predictive value, and negative predictive value of the DDT were evaluated against CCS. Invalidity rates—HPV-negative results with negative internal controls—were compared across sampling methods.

**Results:** The DDT showed a sensitivity of 82.9% (95% CI:72.4–89.9%), specificity of 91.6% (CI:86.4–94.9%), and overall accuracy of 89.0% (CI:84.4–92.4%) relative to CCS. McNemar’s test showed no significant difference between CCS and DDT results (p = 0.845). Valid result rates were highest for DDT (99.2%), followed by VSS (95.4%) and CCS (90.8%).

**Conclusion:** The DDT demonstrates comparable accuracy to CCS for detecting high-risk HPV. This novel device shows promise as a self-sampling method. Further, complementary research should focus on assessing DDT’s clinical performance in detecting HPV associated with cervical disease endpoints.

## INTRODUCTION

Cervical cancer is the fourth most common cancer amongst women worldwide, with an estimated 570,000 new cases and 311,000 deaths annually (1). This preventable cancer is caused by certain types of human papillomavirus (HPV). Cervical cancer screening programs are increasingly based on HPV testing, with a growing trend towards the use of self-sampling methods to identify high-risk HPV. HPV self-sampling can enhance participation rates and provides a less invasive means to engage women than traditional clinician-collected sampling (CSS) methods (2–8).

Studies have demonstrated that self-sampling is cost-effective compared to clinician sampling, with potential cost savings due to reduced need for clinical visits and staff time, supporting its implementation as a viable option in both high-income and low- and middle-income countries (2–4). A randomised study by Aarnio et.al found that self-sampling for HPV testing led to higher participation rates and more cases of high-grade cervical intraepithelial neoplasia (CIN2+) detected at a significantly lower cost compared to clinician based sampling (5). Recent research in the United Kingdom (UK) found that 86% of women preferred having a choice between self-sampling and clinician sampling for cervical screenings, with 69% expressing a preference for self-sampling at home (8).

The WHO have set an ambitious goal to eliminate cervical cancer by the end of the century (9). However, this target faces significant challenges. In the UK, a high income country that offers organised cervical screening, recent data show that only 68.7% of eligible individuals were adequately screened in 2021-2022, well below the 80% target (10). Additionally,iIn low and middle income countries where the majority of cancers manifest, screening programmes are often absent. To achieve the goal of cervical cancer elimination, innovative approaches to screening are urgently needed. These must address both the barriers to participation and the resource limitations within healthcare systems. Whilst vaginal self-sampling (VSS) has been shown to be an effective strategy for increasing cervical cancer screening participation, particularly among underscreened populations (9), some patients also report low self-confidence in obtaining a good quality vaginal sample with self-administered swabs, highlighting the need for more user-friendly self-sampling options (7).

Menstrual tampons could offer a promising alternative for HPV sample collection due to their familiarity (10). Tampons may also have an advantage as screening tools due to their ability to remain in the vaginal canal for an extended period and their larger size compared to other options (11–12). Yet whilst there are several studies which have assessed the analytical and clinical performance of VSS for HPV testing (6), limited research has been conducted on the performance of menstrual tampons as a self-collection device. A systematic review and meta-analysis by Arbyn et al. highlighted the need for further research to validate the use of menstrual tampons for HPV testing (6).

The Daye Diagnostic Tampon (DDT) is an innovative at-home gynaecological test kit that utilises a menstrual tampon for vaginal and cervical sample collection for high-risk HPV testing. Preliminary research has shown that self-collected tampons exhibit accuracy comparable to CCS for detecting HPV and other sexually transmitted infections (13). The present technical study builds on this preliminary data and was designed to compare the performance of the DDT to CSS for the detection of high-risk HPV; the influence of sample collection order on performance and associated comparison(s) was also assessed.

## METHODS

### Study design and setting

This study employed a prospective diagnostic trial design to assess the diagnostic accuracy of the DDT compared to clinician-collected vaginal swabs (CSS) and self-collected vaginal self-swabs (VSS) for the detection of high-risk HPV. As the performance of VSS and DDT may be affected by sampling order, block randomisation was used to assign order of self-sampling with DDT or VSS. Usability and acceptability were determined through questionnaires at baseline and after sampling. To further explore acceptability, participants were offered the opportunity to participate in focus groups after trial completion.

The trial was conducted in the UK between 19th December 2023 and 18th October 2024. Ethical approval was obtained from London Camberwell St Giles Ethics Committee (reference 23-LO-0882). We report against the Standards for Reporting of Diagnostic Accuracy Studies (STARD) guidelines (14). See supplementary 1 for the completed checklist.

### Study population

The study enrolled sexually active individuals assigned female at birth (AFAB) aged 25-65 years. Participants were divided into two groups, with Group 1 consisting of individuals who had received a confirmed HPV-positive screening result within the past four weeks and could provide evidence through the trial ePRO system. All participants were required to be sexually active, defined as having penetrative vaginal sex, and willing to provide informed consent and adhere to trial procedures.

Individuals were ineligible if they had undergone a previous hysterectomy or total hysterectomy with cervix removal, had known allergies or sensitivities to tampons, or had a history of Toxic Shock Syndrome (TSS). The study also excluded pregnant or breastfeeding individuals, those participating in other interventional clinical trials, and anyone using investigational drugs within the previous 30 days.

### Recruitment

Figure 1 depicts the central recruitment flow for the trial. Participants were primarily recruited via social media. Advertising on Meta and other platforms was used to highlight the trial. Between 15th January 2024 - 16th February 2024, adverts directed potential participants to the trial website (see supplementary material 2). If interested in taking part, they completed an online pre-screening form to assess their eligibility for the trial. If potentially eligible, the Participant Information Sheet (PIS) and Informed Consent Form (ICF) were sent directly to the participant. Once enrolled, participants booked an in-person clinic appointment to see a trial nurse at a clinic. Participants self-sampled the day before their in-person clinic appointment. Participants were offered £50 to participate in the trial.

**Figure 1.**
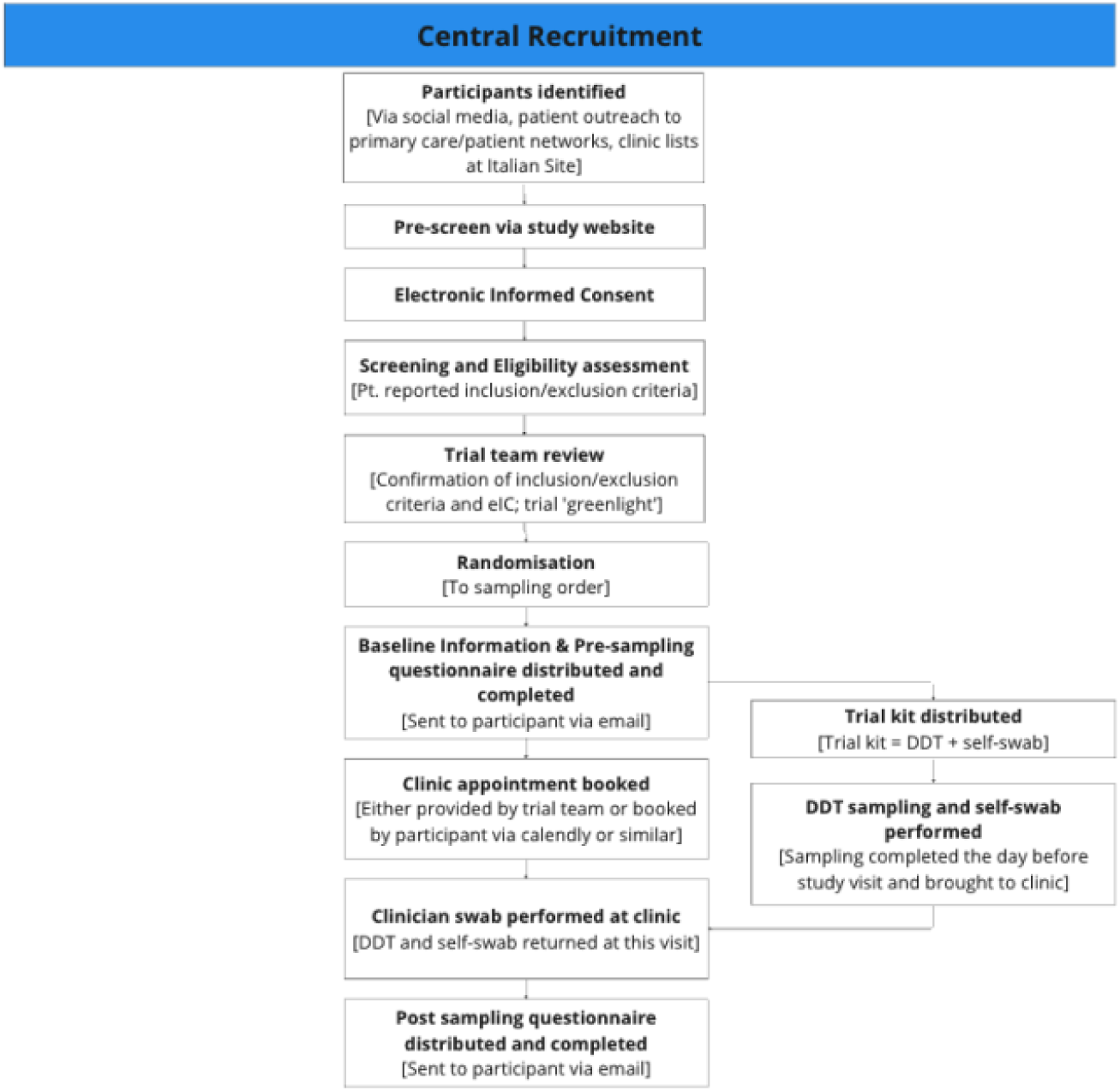
Central Recruitment Flow, showing the participant flow from recruitment from patient outreach (e.g. social media, advertising to patient networks, clinic outreach to patient lists)

### Study procedures and sample collection

Participants underwent pre-screening, informed consent, screening, and eligibility assessments online, where baseline demographic information and medical history were collected.

Participants were then randomly assigned to Group A (self-swab followed by DDT) or Group B (DDT followed by self-swab) using block randomisation. Each participant received a trial kit by post, containing both the DDT and self-swab, along with sterile containers with transport medium, and detailed sampling instructions.

Participants were instructed to use the DDT by inserting it into their vagina, leaving it in place for at least 20 minutes, then removing it before placement in a sterile container with 10 ml of transport medium (Copan UTM). The self-swab was also inserted vaginally and rotated before also being placed into a sterile container with 10ml of transport medium, following the manufacturer’s instructions. Sampling order was specified in the instructions, and participants were asked to allow an hour between the first and second samples.

At a clinic visit the following day, participants provided their self-collected DDT and VSS samples to a nurse, and a clinician-collected vaginal swab was also obtained. After completing all samples, participants filled out additional questionnaires directly in an electronic data capture platform. Participants were offered the opportunity to participate in focus groups, to discuss their experience of using the DDT and preferences for HPV testing. The trial period lasted approximately 2-4 weeks, with all samples sent to an accredited central UK laboratory for analysis.

### Sample Collection Devices

The swabs used for VSS and CSS were Copan FLOQSwabs® which consist of a customisable moulded plastic shaft and a tip coated with perpendicular short Nylon® fibres. Sterile containers for DDT and swab transportation contained 10 ml (DDT) and 3ml (Swab) of transport medium (Copan UTM).

Diagnostic tampons used for self-sampling were sterile, 100% organic cotton, featuring a ’W’ wadding design with a protective sleeve and a bio-based polyethylene applicator (Figure 2). The applicator ensures the patient can collect a sample from the cervical area, as well as from the vaginal canal. A withdrawal cord, made from mercerized organic cotton, is attached to the pledget.

**Figure 2.**
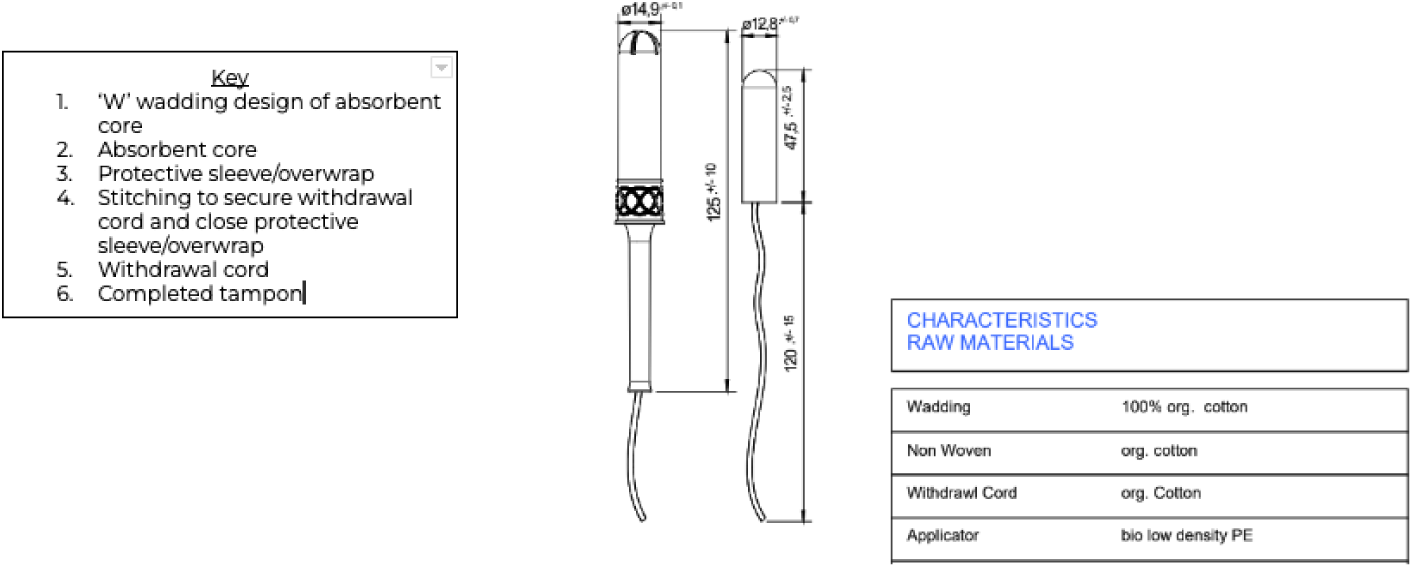
Structure, dimensions, and characteristics of the tampon as well as its raw materials.

### HPV testing

The Aptima® HPV assay was utilised for HPV detection. This assay targets the expression of E6/E7 mRNA of 14 high-risk HPV types (16, 18, 31, 33, 35, 39, 45, 51, 52, 56, 58, 59, 66, and 68). While the assay was performed according to manufacturer’s instructions the bio-samples used in the evaluation were not formally validated by the manufacturer;

### Reference and index sampling methods

In this study, the gold standard or reference sampling method used was CCS, as used in many other self-sampling diagnostic accuracy studies. Our primary index sampling method of investigation was the DDT, however we also calculated the accuracy of VSS as an alternate index. As an exploratory sensitivity analysis, we also explored a collated measure of results from CCS, VSS and DDT as an alternative reference sampling method. With this collated measure, a positive result was recorded only when at least two sampling methods showed positive results. If only one of the three methods showed a positive result, the outcome was considered inconclusive. A negative result required all three samples to test negative.

### Statistical analyses

Diagnostic accuracy parameters (sensitivity, specificity, overall accuracy, positive and negative predictive values) were calculated from all complete and conclusive results and reported with Wilson score 95% confidence intervals. Invalidity rates—HPV-negative results with negative internal controls—were compared across sampling methods.

Additional sensitivity analyses were conducted. To compensate for variation in conclusive results, diagnostic accuracy parameters were repeated using the collated measure as the gold standard. McNemar tests were conducted to assess whether diagnostic accuracy differed significantly between CCS and collated results as gold standards. To understand the impact of sampling order, a sensitivity analysis was conducted assessing diagnostic accuracy for tampon swabs and self-swabs when the respective sampling method was used first vs. second. McNemar tests were conducted to compare diagnostic accuracy between DDT vs. VSS first orders for VSS and DDT, respectively.

### Sample size

A sample size of 420 participants was calculated to provide a statistical power of 90% to test whether the sensitivity of DDT is above 70% when the assumed true sensitivity is 95% and the prevalence of HPV in the recruited patient population is 5%. This was assumed to yield 21 cases of HPV to evaluate the sensitivity of HPV.

### Focus groups

Focus groups were held online via Google meet, a video calling platform. Participants invited to participate via email following their completion of their clinic visit. Participants were offered an additional £25 to take part in these groups. Informed consent was taken before participation. The groups were facilitated by VM and MG, with support from another facilitator (MT). The topic guide can be found in supplementary material (supplementary 3).

## RESULTS

### Sample

From an initial screening of 489 participants, 437 were enrolled, with 52 screen failures. Of the 329 booked in for a clinic visit, 263 participants attended a clinic visit for CSS, having completed DDT and VSS and all questionnaires for acceptability outcomes. As three participants either did not complete CSS in clinic or had missing CSS results, the final sample used for diagnostic accuracy results consisted of 260 participants who successfully completed all study requirements, with results from all three sampling methods (CSS, VSS and DDT). In the final diagnostic accuracy sample, 26 participants had a confirmed HPV+ screening result within the previous 4 weeks (Group 1). See figure three for the participant flow chart.

**Figure 3.**
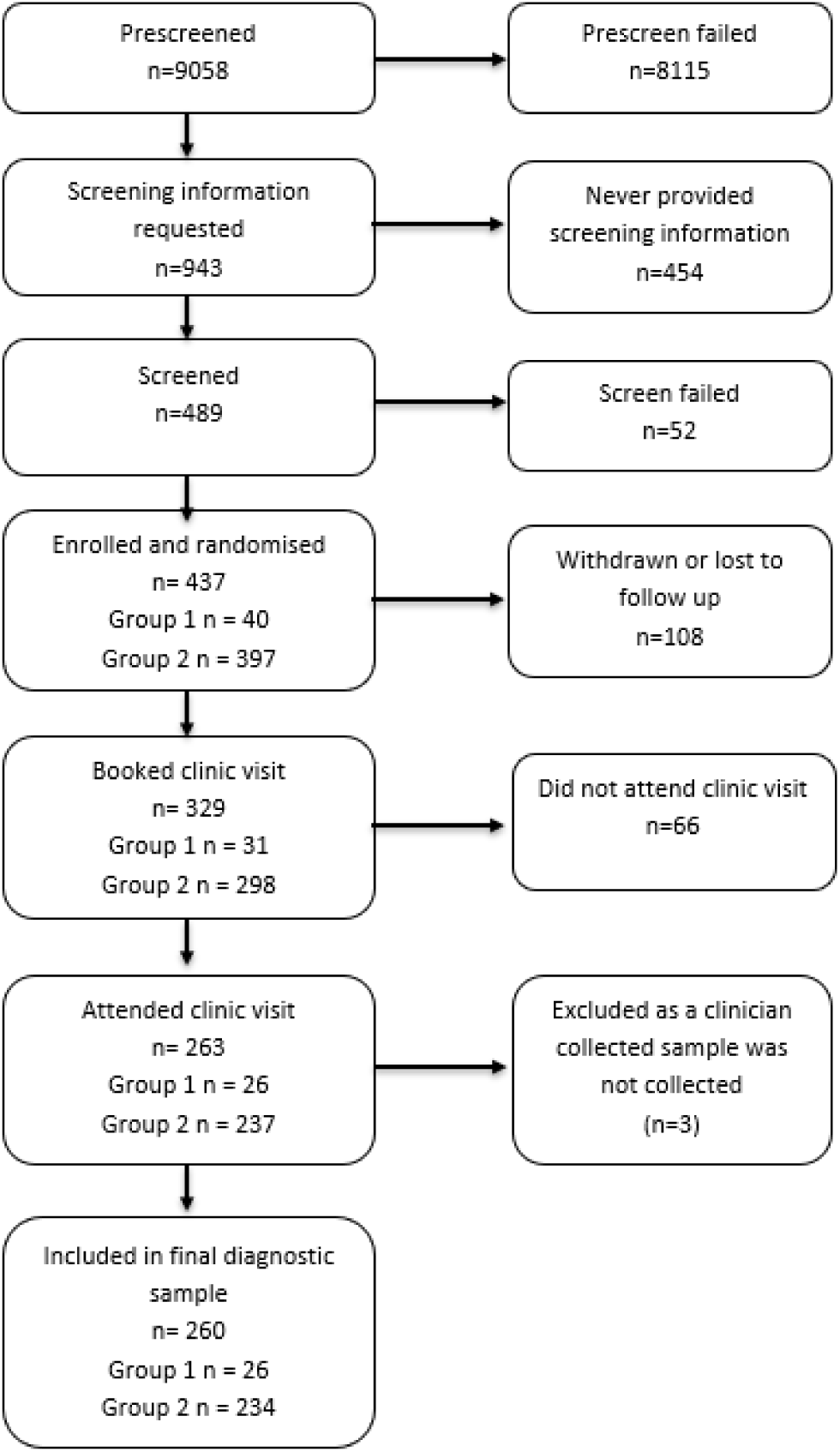
Participant flowchart.

### Participant Characteristics

The median age was 31.0 years, with an interquartile range between 28.0 and 36.0 years. Most participants were white (69%), heterosexual (75%), and single (59%, see Table 1). Characteristics of the sample used for diagnostic accuracy results (n=260), as a whole and broken down by randomisation arm, can be found in supplementary material 4, table 1.

**Table 1.** Participant characteristics.

| Characteristic | N = 263<sup>1</sup> |
| --- | --- |
| Age | 31 (28, 36) |
| **Ethnicity** |  |
| Asian | 16 (6.1%) |
| Black, Caribbean or African | 25 (9.5%) |
| Other | 39 (14.8%) |
| White | 183 (69.6%) |
| **Sexual Orientation** |  |
| Heterosexual | 197 (74.9%) |
| Bisexual | 49 (18.6%) |
| Prefer not to say | 7 (2.7%) |
| Lesbian/Gay | 6 (2.3%) |
| Other | 4 (1.5%) |
| **Relationship Status** |  |
| Single | 155 (58.9%) |
| Co-habiting | 53 (20.2%) |
| Married | 27 (10.3%) |
| Civil Partnership | 12 (4.6%) |
| Prefer not to say | 10 (3.8%) |
| Divorced | 6 (2.3%) |

<sup>1</sup>Median (IQR); n (%)

### Diagnostic Accuracy: CSS as the gold standard

A total of 780 samples (one sample per sampling method) from 260 participants were analysed. As the primary outcome, diagnostic accuracy was determined using CCS as the gold standard.

For the CCS, 70 participants tested positive for HPV (26.7%) 168 participants tested negative (64.6%). For 22 participants, the test result was invalid (8.5%). Of the 22 samples that were invalid with the CCS, 21 were evaluated as positive (n = 3) or negative (n = 18) using the DDT, with 1 sample also returning an invalid result. DDT produced 2 invalid sample results, 1 of which was evaluated as negative in CCS, the other as invalid (see supplementary material 5, Table 2). Comparing results from VSS, 5 of the 22 invalid CCS results were also evaluated as invalid, while 14 were assessed as negative and 3 as positive for HPV. An additional 9 samples yielded invalid results in VSS ((See supplementary material 6, Table 3).

Diagnostic accuracy metrics for both DDT and VSS vs CCS are reported in Table 2. Using the CCS as gold standard yields a sensitivity of DDT of 82.9% (95% CI: 72.4% - 89.9%), specificity of 91.6% (86.4 % - 94.9%), PPV of 80.6% (70.0% - 88.0%), NPV of 92.7% (87.7% - 95.8%), and overall accuracy of 89.0% (84.4% - 92.4%). McNemar’s tests did not reveal a significant deviation of CCS and DTT results (χ^2^= 0.04, df = 1, *p* = 0.845), nor of CCS and VSS results (χ^2^ = 0.03, df = 1, *p* = 0.860), suggesting results do not differ in terms of sensitivity and specificity.

**Table 2:** Diagnostic accuracy of DDT and VSS compared to CCS.

|  | Daye Diagnostic Tampon (DDT)<sup>1</sup> | Vaginal self-sampling (VSS)<sup>1</sup> |
| --- | --- | --- |
| Sensitivity | 82.9 (72.4 – 89.9) | 75.4 (64.0 – 84.0) |
| Specificity | 91.6 (86.4 – 94.9) | 90.6 (85.1 – 94.2) |
| PPV | 80.6 (70.0 – 88.0) | 77.6 (66.3 – 85.9) |
| NPV | 92.7 (87.7 – 95.8) | 89.5 (83.8 – 93.3) |
| Accuracy | 89.0 (84.4 – 92.4) | 86.0 (80.9 – 89.9) |
| <sup>1</sup> All values in percent. Values in parentheses indicate 95% CI limits |  |  |

### Sensitivity analysis: Using collated results as the gold standard

The proportion of conclusive results varied between the three sampling methods – CCS yielded results in 238 samples (90.8%), DDT in 258 (99.2%), and VSS in 248 (95.4%) out of 260 samples. Therefore, as an exploratory sensitivity analysis, we explored using a collated measure as the gold standard.

Using the collated results as a reference, 67 participants were classified as positive for HPV (25.8%), 178 participants were categorised as negative (68.4%), and for 2 participants, the test result was invalid (0.8%), inconclusive (n = 14, 5.4%), or missing (n = 1, 0.4%).

CCS detected 62 of the 67 HPV-positive cases and 156 of the 178 HPV-negative samples (see supplementary 7, Table 4). Compared to the collated as a reference, CSS yielded a sensitivity of 95.4% (87.3% - 98.4%), specificity of 96.3% (92.2% - 98.3%), PPV of 91.2% (82.1% - 95.9%), NPV of 98.1% (94.6% - 99.4%), and overall accuracy of 96.0% (92.6% - 97.9%) [see table 3]. A McNemar’s test did not reveal a significant deviation of CCS and collated results (χ^2^ = 0.44, df = 1, *p* = 0.504).

**Table 3:** Diagnostic accuracy of CCS, DDT and VSS compared to collated results as reference.

|  | Clinician-collected Swabs (CSS)<sup>1</sup> | Daye Diagnostic Tampon (DDT)<sup>1</sup> | Vaginal self-sampling (VSS)<sup>1</sup> |
| --- | --- | --- | --- |
| Sensitivity | 95.4 (87.3 – 98.4) | 92.5 (83.7 – 96.8) | 84.6 (73.9 – 91.4) |
| Specificity | 96.3 (92.2 – 98.3) | 96.0 (91.9 – 98.1) | 94.6 (90.1 – 97.1) |
| PPV | 91.2 (82.1 – 95.9) | 89.9 (80.5 – 95.0) | 85.9 (75.4 – 92.4) |
| NPV | 98.1 (94.6 – 99.4) | 97.1 (93.4 – 98.8) | 94.0 (89.4 – 96.7) |
| Accuracy | 96.0 (92.6 – 97.9) | 95.0 (91.5 – 97.1) | 91.8 (87.6 – 94.7) |
| <sup>1</sup> All values in percent. Values in parentheses indicate 95% CI limits |  |  |  |

Using DDT as a sampling method yielded correct positive results in 62 of 67 participants categorised as HPV-positive, and correct negative results in 167 out of 176 HPV-negative participants, when using the collated measure as a reference (see supplementary 8, Table 5). DDT sensitivity was 92.5% (95% CI: 83.7% - 96.8%), specificity 96.0% (91.9% - 98.1%), PPV 89.9% (80.5% - 95.0%), NPV 97.1% (93.4% - 98.8%), and overall accuracy 95.0% (91.5% - 97.1%), see Table 3. A McNemar’s test did not reveal a significant deviation of DDT and collated results (χ^2^ = 0.08, df = 1, *p* = 0.773).For VSS, diagnostic accuracy metrics are reported in supplementary material 9, Table 6. A McNemar’s test did also not indicate a significant deviation of VSS and collated results (χ^2^ < 0.01, df = 1, *p* > 0.999).

### Sensitivity analysis: Sampling order

#### Vaginal self-sampling (VVS)

Sensitivity of VSS was higher when sampled first vs. second (88.9% vs. 79.3%, χ2 = 16.45, p < 0.001). See Table 4. Specificity was slightly but significantly higher when DDT were sampled first compared to VSS first (self-swabs first: 93.3%, DDT first: 95.7%, χ2 = 57.10, p < 0.001).

**Table 4:** Diagnostic accuracy for VSS by sampling order.

| | VSS diagnostic accuracy when VSS sampled first<sup>1</sup> | VSS diagnostic accuracy when DDT sampled first<sup>1</sup> | $\chi^2$ | $p$ |
| --- | --- | --- | --- | --- |
| Sensitivity | 88.9 (74.7 – 95.6) | 79.3 (61.6 – 90.2) | 16.45 | < 0.001 |
| Specificity | 93.3 (85.3 – 97.1) | 95.7 (89.3 – 98.3) | 57.10 | < 0.001 |
| PPV | 86.5 (72.0 – 94.1) | 85.2 (67.5 – 94.1) |  |  |
| NPV | 94.6 (86.9 – 97.9) | 93.6 (86.8 – 97.0) |  |  |
| Accuracy | 91.9 (85.3 – 95.7) | 91.7 (85.5 – 95.4) |  |  |
| <sup>1</sup> All values in percent. Values in parentheses indicate 95% CI limits |  |  |  |  |

#### Daye diagnostic tampon (DDT)

For DDT, sensitivity and specificity improved significantly when DDT were sampled first vs. second (sensitivity: 100% vs. 86.5%, χ2 = 30.03, p < 0.001; specificity: 96.8% vs. 95.0%, χ2 = 65.62, p< 0.001). See Table 5.

**Table 5:** Diagnostic accuracy for tampon swabs by sampling order.

| | DDT diagnostic accuracy when DDT sampled first<sup>1</sup> | DDT diagnostic accuracy when VSS sampled first<sup>1</sup> | $\chi^2$ | $p$ |
| --- | --- | --- | --- | --- |
| Sensitivity | 100 (88.6 – 100) | 86.5 (72.0 – 94.1 ) | 30.03 | < 0.001 |
| Specificity | 96.8 (91.0 – 98.9) | 95.0 (87.8 – 98.0) | 65.62 | < 0.001 |
| PPV | 90.9 (76.4 – 96.9) | 88.9 (74.7 – 95.6) |  |  |
| NPV | 100 (95.9 – 100) | 93.8 (86.4 – 97.3) |  |  |
| Accuracy | 97.6 (93.1 – 99.2) | 92.3 (86.0 – 95.9) |  |  |
| <sup>1</sup> All values in percent. Values in parentheses indicate 95% CI limits |  |  |  |  |

### Acceptability

#### Questionnaire data

Participant questionnaire responses (n=263) revealed high acceptance of tampon-based testing HPV. Prior to sampling, 98.5% (259/263) had used tampons, but only 29.7% (78/263) were aware of their potential for diagnostic testing. Post-sampling, tampons became the preferred testing method (46%), followed by self-swabs (33%). Comfort levels with tampon use for testing remained high (Very comfortable: 206/263, 78.3% pre-sampling; 192/263 73.0% post-sampling), and perceived ease of use increased from 63.5% (167/263) to 74.5% (196/263). While moderate or extreme concerns about accuracy slightly increased post-sampling (10.2% [27/263] to 13.6% [36/263], p < 0.05), overall acceptance remained strong. Trust in tampon results compared to clinician swabs remained stable.

**Table 9.** Acceptability results.

|  | Pre-sampling<br>questionnaire<br>(n=263), n (%) | Post-sampling<br>questionnaire<br>(n=263), n (%) |
| --- | --- | --- |
| Tampon use prior to trial | 259 (98.5) | - |
| Aware tampons could be used for testing for HPV, STIs, or BV | 78 (29.7) | - |
| Level of comfort with idea of using a tampon for sample collection |  |  |
| Very comfortable | 206 (78.3) | 192 (73.0) |
| Comfortable | 51 (19.4) | 49 (18.6) |
| Neither comfortable nor uncomfortable | 6 (2.3) | 5 (1.9) |
| Uncomfortable | - | 5 (1.9) |
| Very uncomfortable | - | 12 (4.5) |
| Perceived ease of tampon use for HPV sample collection |  |  |
| Very Easy | 167 (63.5) | 196 (74.5) |
| Easy | 84 (31.9) | 54 (20.5) |
| Neither easy nor difficult | 7 (2.7) | 8 (3.0) |
| Difficult | - | 3 (1.1) |
| Very difficult | 5 (1.9) | 2 (0.7) |
| Concerns about accuracy |  |  |
| Not at all concerned | 69 (26.2) | 63 (23.9) |
| Slightly concerned | 98 (37.3) | 115 (43.7) |
| Somewhat concerned | 69 (26.2) | 49 (18.6) |
| Moderately concerned | 19 (7.2) | 25 (9.5) |
| Extremely concerned | 8 (3.0) | 11 (4.1) |
| Trust in tampon results compared to clinician swabs |  |  |
| Equal trust in tampon and clinician swabs | 151 (57.4) | 144 (54.7) |
| Greater trust in clinician swab | 106 (40.3) | 114 (43.3) |
| Greater trust in tampon | 6 (2.3) | 5 (1.9) |

#### Focus groups

Four focus groups were held with a total of 24 participants (5-7 participants in each group). Sessions lasted 60 minutes.

Of the 17 unique participants who stated preferences for HPV sampling methods during focus group discussions, 12 (70.5%) shared a strong preference for DDT. Four participants held more neutral positions, sharing mixed views and preferences between DDT, VSS and visiting a clinic for sampling. One participant strongly preferred using clinic services. The participant who preferred clinic-based testing related this to their concerns about the accuracy and comfort of the DDT. Two other participants also had initial concerns about the accuracy of the DDT, despite their preference for it over other sampling methods. Other concerns raised in the group related to the convenience (1), cost (1) or sustainability (1) of the DDT, or to the clarity of information provided around self-sampling with the DDT.

More extensive acceptability results from questionnaires and focus groups will be reported in a future publication.

## DISCUSSION

The primary objective of this study was to assess the accuracy of the DDT for detecting HPV infection. We found that, when using CCS as the reference standard, DDT exhibited a sensitivity of 82.9%, specificity of 91.6%, and overall accuracy of 89.0%. DDT diagnostic performance was as good as that of VSS, which had a sensitivity of 75.4%, a specificity of 90.6% and an overall accuracy of 86.0% when compared with CSS. Results from questionnaires and focus groups suggest that tampons are an acceptable method of self-sampling for HPV, although comfort and perceived accuracy may be concerns for some.

The findings of this study align with the vast body of literature that exists about the comparable accuracy of VSS vs CCS (15,16). A meta-analysis of six comparable studies to ours, where VSS sought to identify the presence of HPV, rather than of precancerous cell changes (CIN2+), found a pooled sensitivity of 74% (61-84%) and specificity of 88% (83- 92%), when compared to CCS (15). Limited published evidence exists on the diagnostic performance of tampons as self-sampling devices for high-risk HPV. We know of two relevant studies in South Africa which assessed the acceptability and accuracy of tampon-based self-collection for high-risk HPV testing, compared to CSS, one study with women living with HIV (17), another with gynaecology clinic attendees (18). Tampon self-sampling in Adamsnon’s study exhibited a sensitivity of 77% and specificity of 77% for detecting high-risk HPV, whilst the comparable sensitivity and specificity in Tito’s study were 86% and 88%, respectively. Unlike another older study of a different device which found tampons to have a high proportion of insufficient quality specimens (19), the DDT only yielded 0.8% (2/260) invalid results, when compared to CSS results.

We found the DDT to be an acceptable method for HPV self-sampling. This echoes the few other UK-based and global studies conducted, which have found it to be an acceptable, convenient and easy to use option (17,18, 20). Though potentially a more supportive sample, a study in London (n=501) where women were recruited from colposcopy units, found the majority of participants (98%) approved of tampons being used as a self-collection tool to test for HPV infection (20). The above-mentioned South African studies also reported high levels of acceptability. Adamson’s study of women living with HIV (n=325) found that over 90% of participants reported no difficulties in collecting HPV samples with a tampon, and 82% were willing to self-collect with a tampon at home (17). Another study in South Africa (n=527) found very similar results showing self-sampling with a tampon to be a comfortable, painless option, preferable for many (18).

Sensitivity analyses showed that the diagnostic performance of the DDT increased to a sensitivity of 92.5% (83.7 – 96.8), a specificity of 96.0% (91.9 – 98.1) and an overall accuracy of 95.0% (91.5 – 97.1) when using the collated measure as the reference standard, rather than CCS. We conducted the sensitivity analysis with a collated measure to mitigate against solely relying on the CCS given that it produced the highest invalidity rate. We suspect that the robustness of CSS in our study is limited by the fact that the copan swab used for both clinician and self collection is not formally approved for use for APTIMA testing. As the APTIMA HPV test used does not include an endogenous cellular control, invalidity was driven by other reasons beyond a lack of available cellular material which likely included inhibition. The novel approach of using a composite measure leverages all available information and mitigates against the high invalidity rate from CSS alone. Notably, DDT and CCS performed very similarly when using the collated results.

Sensitivity analyses also suggest that sampling order may have had an impact on DDT’s diagnostic performance. The analysis of sampling order revealed significant effects of the order of sampling on diagnostic accuracy for VSS and DDT. Sensitivity increased for both methods when sampled first, which is consistent with expectation and published evidence. Specificity decreased slightly (but significantly) for VSS when the VSS was sampled first; interestingly, specificity was increased when the DDT was taken first. Reasons for the specificity observations are not entirely clear; whilst the dimensions and absorbency of the different devices are likely to play a role, these data underline the importance of the evaluation and validation of individual devices.

### Study strengths and limitations

This is a timely study of an innovative device that could broaden choice within HPV self-sampling (9–10). It is important to note the contribution this study makes to a gap in evidence, as there are limited data on the acceptability and performance of tampons as biospecimens for HPV testing, in particular where the impact of sampling order is assessed (17,18,20). Consequently, we hope these data act as a grounding for further development and evaluation of the DDT.

Some limitations should be considered. We experienced greater loss to follow up than anticipated, resulting in a smaller sample than planned of 260. However, due to targeted recruitment of those with a confirmed HPV result, which enriched our population, we superseded the required number of cases to confidently evaluate the DDT sensitivity if greater than 70%, compared to CSS. In this study, we only used one HPV assay; further work could enhance findings by evaluating other DDT and HPV assay combinations, increasing reliability. As mentioned, despite being the commonly referenced gold-standard, CCS produced the highest invalidity rate amongst the three sampling methods. To mitigate against this, we conducted a sensitivity analysis with a collated measure, leveraging all available results. Most importantly, we recognize that our assessment focused on diagnostic accuracy for HPV detection, rather than HPV associated with histologically confirmed disease, specifically CIN2+. The absence of follow-up limits our ability to assess the sampling methods’ longitudinal performance (21). To address this, ongoing work is evaluating the clinical performance of the DDT relative to CIN2+, which will naturally extend the data presented here. Additionally, it is crucial to assess the effectiveness of DDT in diverse populations, considering various age groups, ethnicities, and socioeconomic backgrounds, to ensure broad applicability and address potential confounding factors (22). Whilst we did achieve a diverse sample in terms of ethnicity and sexuality, which is of particular importance due to lower screening in these groups (23, 24) , future studies could be repeated with older participants. This is particularly important given that cervical cancer screening programmes in the UK suggest screening until women and individuals AFAB reach 64-years-old (25).

## Conclusion

This study confirms the value of diagnostic tampons as an alternate method to CCS for detecting high-risk HPV infection, with high levels of diagnostic accuracy. Notably, DDT performed similarly to CCS when using the collated results as the reference standard. The performance and ease of use of DDT highlight its potential as a valuable self-collection method for cervical cancer screening. Further research is needed to validate these findings in longitudinal cohorts and relative to clinical endpoints. Understanding practical challenges and logistical considerations of DDT implementation, as well as comprehensive cost-effectiveness analyses, will help determine the financial viability of integrating DDT into screening programs, particularly in low-income countries (26).

## Acknowledgements

This study was made possible through the valuable contributions of Lindus Health for their excellent organisation of the clinical trial, Alderley Lighthouse Labs for their crucial lab work, and Dr. Kate Cuschieri from the University of Edinburgh for her collaboration, scientific consultation, and lab work. The authors express their deepest gratitude to these partners for their instrumental roles in successfully completing this impactful research project. Thank you to Mia Taicher for their support facilitating focus groups.

## Data availability

Both the full study protocol and the study data are available on reasonable request, with requests made to the corresponding author.

## Ethical Approval

Ethical approval was obtained from London Camberwell St Giles Ethics Committee (reference 23-LO-0882).

## Supplementary material

### Supplementary material 1. STARD checklist

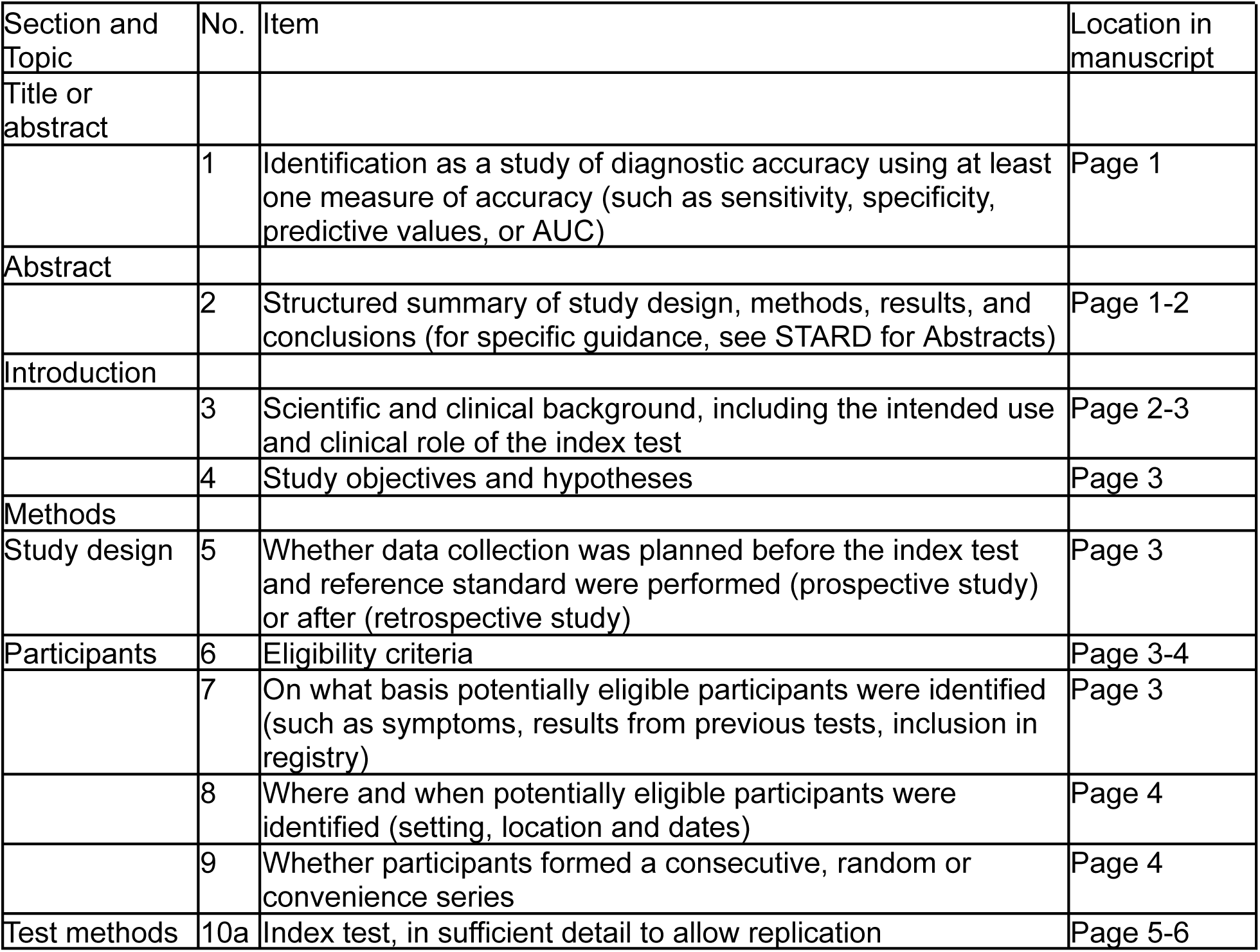
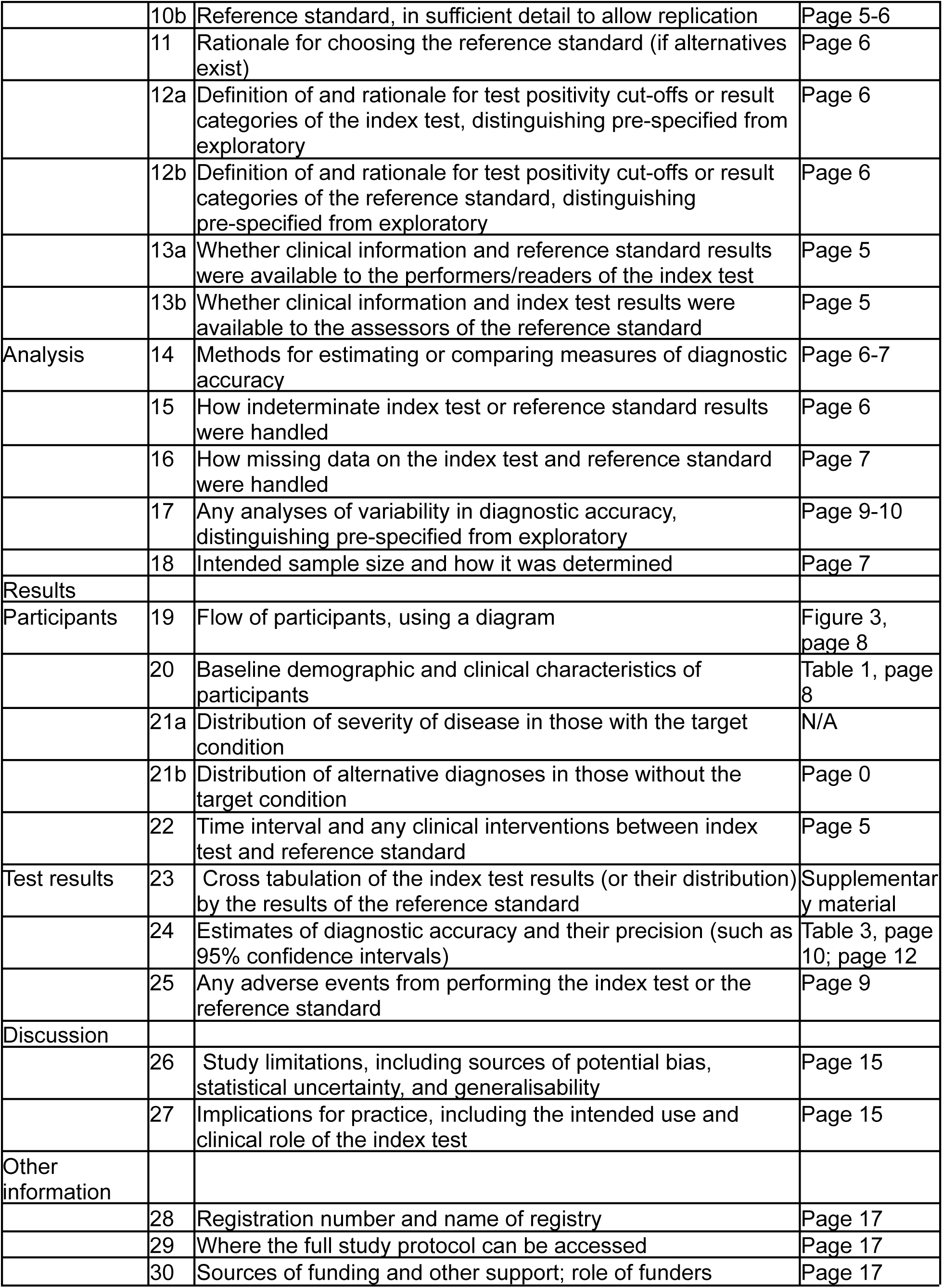

Bossuyt, P.M., Reitsma, J.B., Bruns, D.E., Gatsonis, C.A., Glasziou, P.P., Irwig, L., et al. (2015). STARD 2015: An updated list of essential items for reporting diagnostic accuracy studies

### Supplementary material 2. Social media recruitment adverts

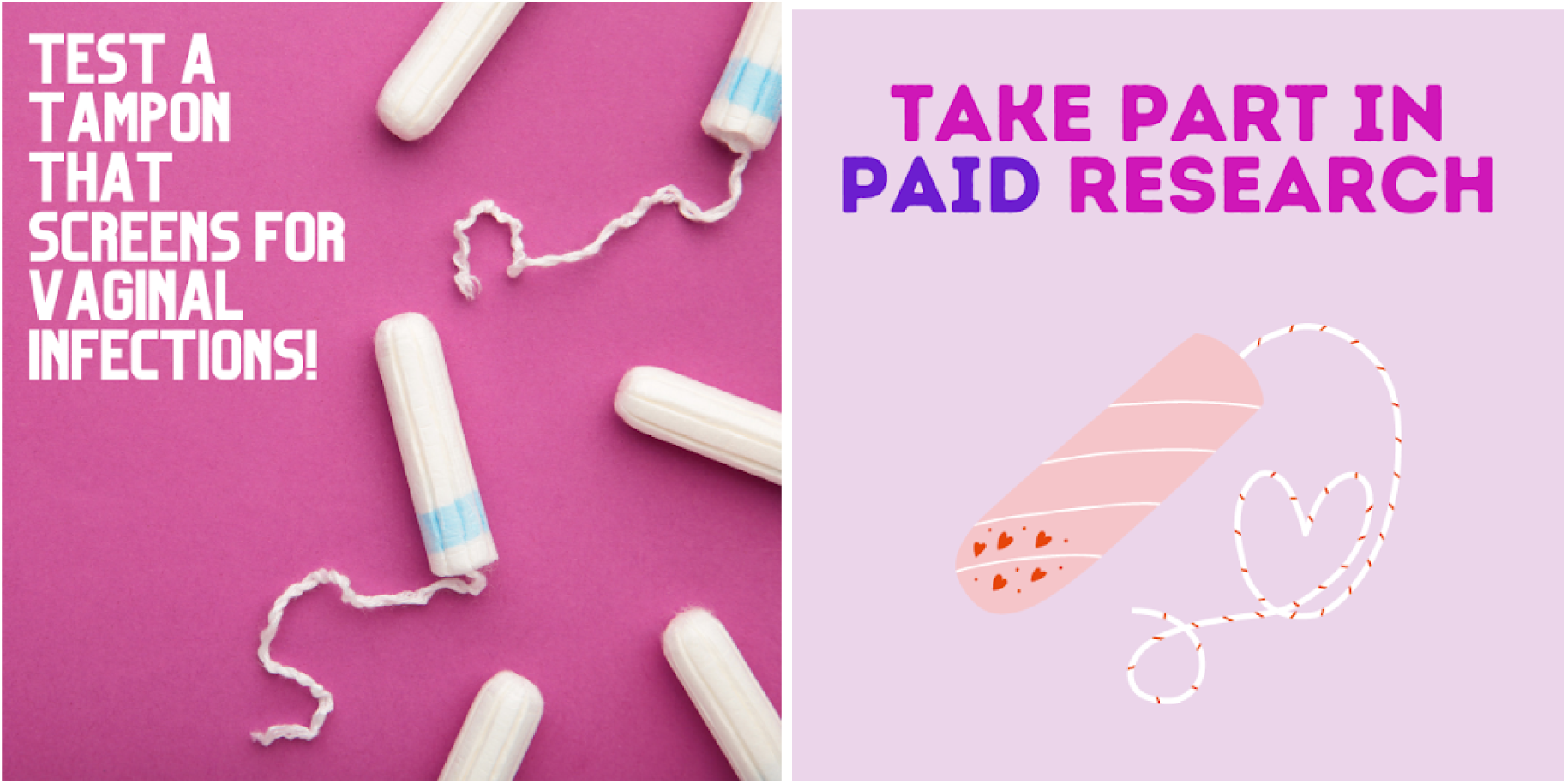

### Supplementary material 3. Focus group topic guide

1. Motivation for Joining:

- What motivated you to participate in the STAMP trial?
- What is your previous experience with HPV testing?
- What was your experience with the communication and logistics surrounding the trial?
2. Experience with Collection Methods:

- Can you describe your experience with the self-collected tampon? Compared to the other
- two methods
- are you familiar with tampons already? Applicator vs non applicator?
- What challenges did you face during the sample collection process?
3. Usability of the DT:

- How did you find the instructions for using the tampon?
- do you have experience with at home testing kits?
- what are your perceptions of at home testing kits?
- did you have any concerns about doing the self sampling yourself?
- What improvements would you suggest for the sample collection process with the DT?
4. Comfort Using the DT:

- How comfortable were you using the tampon for sample collection?
- What could be done to improve your comfort level?
- Did you feel anxious prior to sample collection?
5. Acceptability and Perceptions:

- How confident do you feel about using a tampon for sample collection in terms of
- accuracy?
- If the tampon was offered by the NHS, would you be more confident about it?
- Which sample collection method would you prefer for future tests? Factors that influence
- Would you recommend the tampon-based sampling to others? Why or why not?

## Supplementary material 4

**Table 1.**
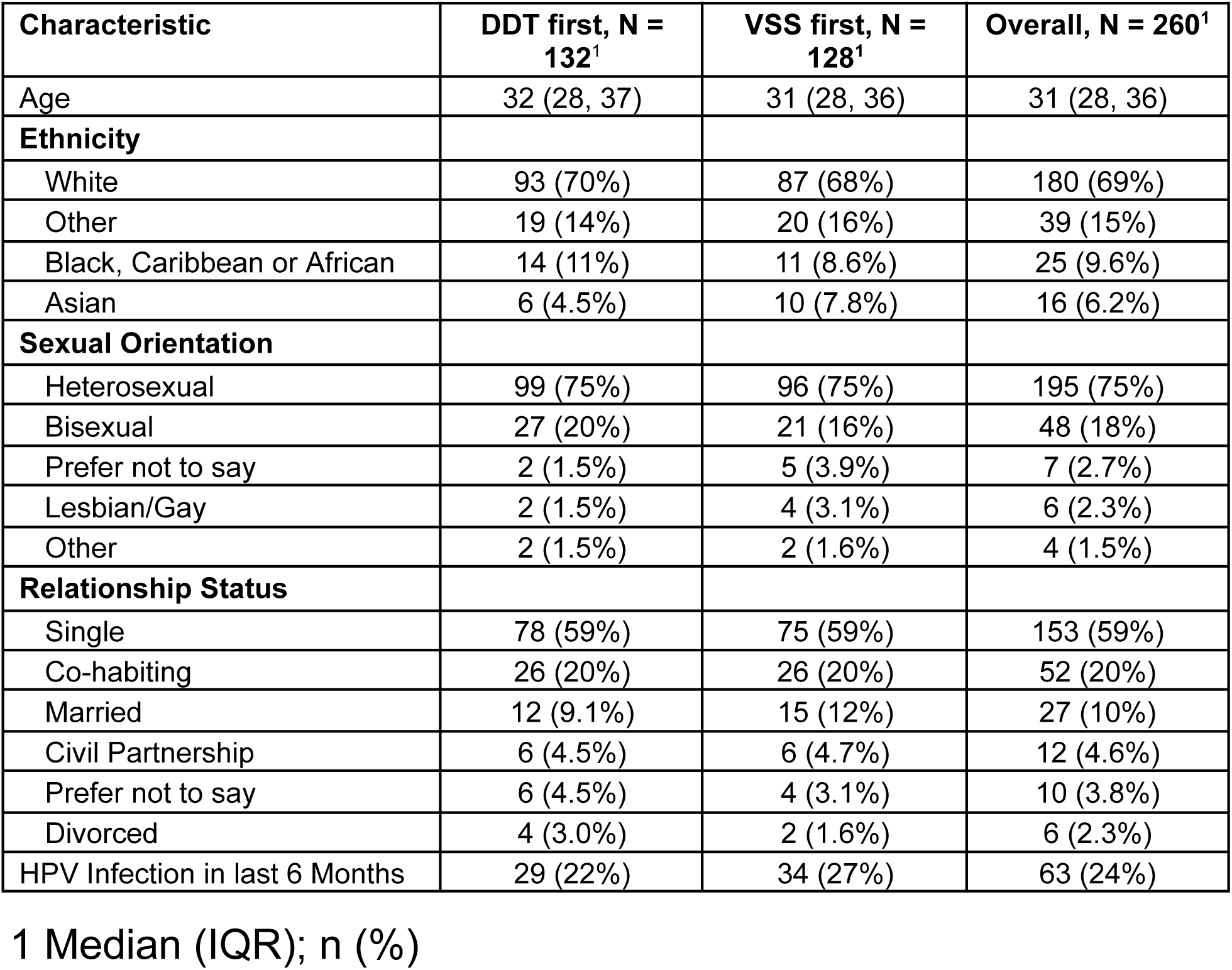
Participant characteristics of diagnostic accuracy analyses (n=260), as a whole and by randomisation arm.

| Characteristic | DDT first, N = 132<sup>1</sup> | VSS first, N = 128<sup>1</sup> | Overall, N = 260<sup>1</sup> |
| --- | --- | --- | --- |
| Age | 32 (28, 37) | 31 (28, 36) | 31 (28, 36) |
| **Ethnicity** |  |  |  |
| White | 93 (70%) | 87 (68%) | 180 (69%) |
| Other | 19 (14%) | 20 (16%) | 39 (15%) |
| Black, Caribbean or African | 14 (11%) | 11 (8.6%) | 25 (9.6%) |
| Asian | 6 (4.5%) | 10 (7.8%) | 16 (6.2%) |
| **Sexual Orientation** |  |  |  |
| Heterosexual | 99 (75%) | 96 (75%) | 195 (75%) |
| Bisexual | 27 (20%) | 21 (16%) | 48 (18%) |
| Prefer not to say | 2 (1.5%) | 5 (3.9%) | 7 (2.7%) |
| Lesbian/Gay | 2 (1.5%) | 4 (3.1%) | 6 (2.3%) |
| Other | 2 (1.5%) | 2 (1.6%) | 4 (1.5%) |
| **Relationship Status** |  |  |  |
| Single | 78 (59%) | 75 (59%) | 153 (59%) |
| Co-habiting | 26 (20%) | 26 (20%) | 52 (20%) |
| Married | 12 (9.1%) | 15 (12%) | 27 (10%) |
| Civil Partnership | 6 (4.5%) | 6 (4.7%) | 12 (4.6%) |
| Prefer not to say | 6 (4.5%) | 4 (3.1%) | 10 (3.8%) |
| Divorced | 4 (3.0%) | 2 (1.6%) | 6 (2.3%) |
| HPV Infection in last 6 Months | 29 (22%) | 34 (27%) | 63 (24%) |

1 Median (IQR); n (%)

## Supplementary material 5

**Table 2:** DDT vs. CCS results.

|  |  | Daye Diagnostic Tampon (Index Result) |  |  |  |
| --- | --- | --- | --- | --- | --- |
|  |  | Positive | Negative | Invalid | Total |
| Clinician collected sampling (Reference Result) | Positive | 58 | 12 | 0 | 70 |
|  | Negative | 14 | 153 | 1 | 168 |
|  | Invalid | 3 | 18 | 1 | 22 |
|  | Total | 75 | 185 | 2 | 260 |

## Supplementary material 6

**Table 3:** VSS vs. CCS results.

|  |  | Vaginal Self-Swab (Index Result) |  |  |  |
| --- | --- | --- | --- | --- | --- |
|  |  | Positive | Negative | Invalid | Total |
| Clinician collected sampling (Reference Result) | Positive | 52 | 17 | 1 | 70 |
|  | Negative | 15 | 145 | 8 | 168 |
|  | Invalid | 3 | 14 | 5 | 22 |
|  | Total | 76 | 178 | 14 | 260 |

## Supplementary material 7

**Table 4:**
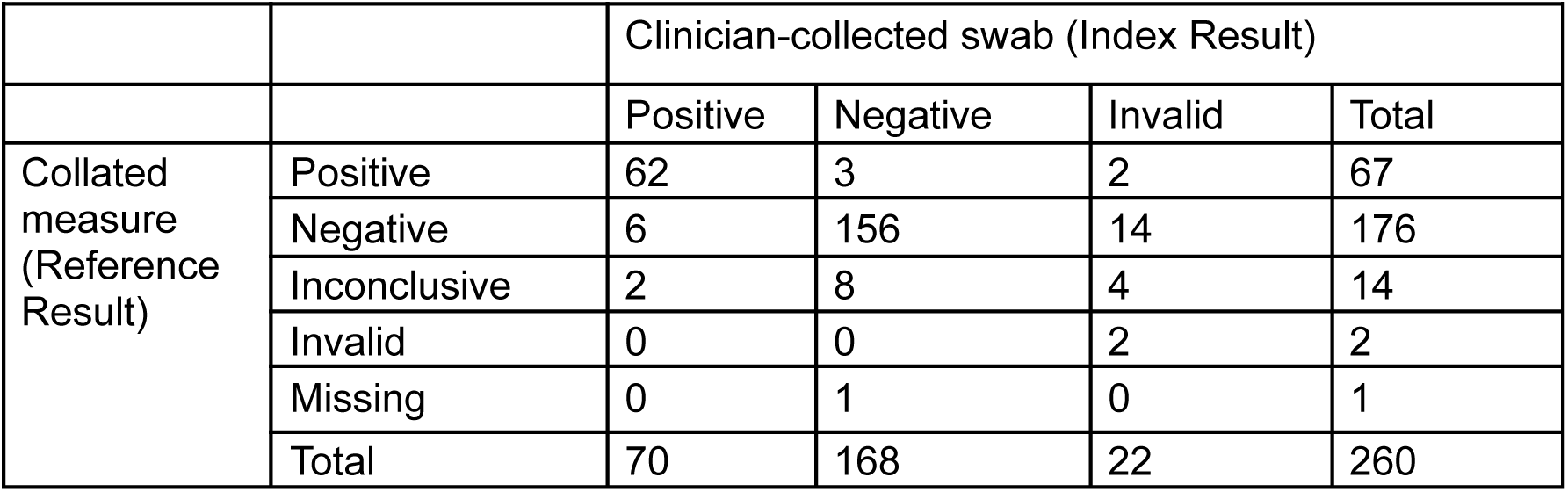
CCS vs. collated results.

## Supplementary material 8

**Table 5:**
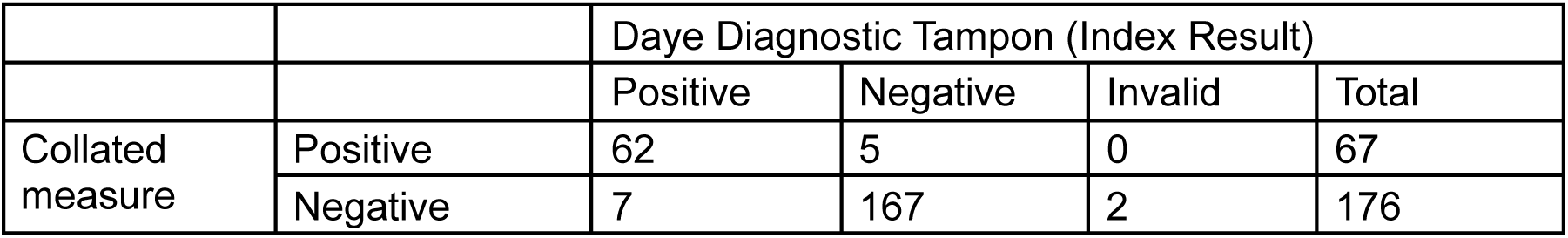
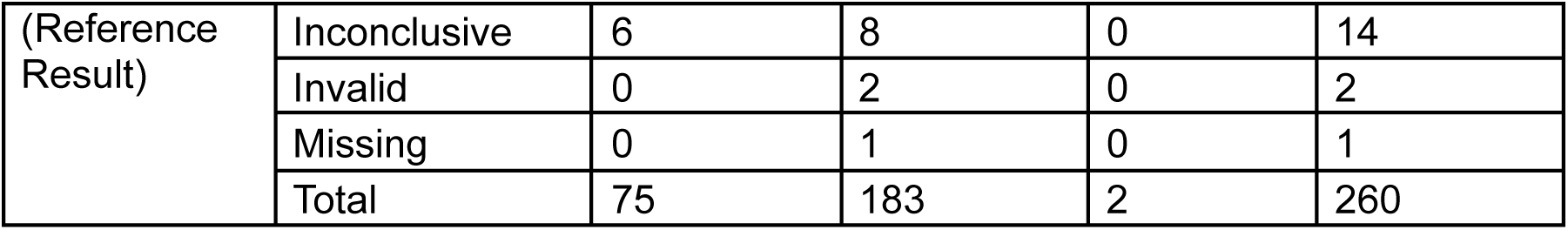
DDT vs. collated results.

## Supplementary material 9

**Table 6:** VSS vs. collated results.

|  |  | Vaginal Self-Swab (Index Result) |  |  |  |
| --- | --- | --- | --- | --- | --- |
|  |  | Positive | Negative | Invalid | Total |
| Collated measure (Reference Result) | Positive | 55 | 10 | 2 | 67 |
|  | Negative | 9 | 158 | 9 | 176 |
|  | Invalid | 0 | 0 | 2 | 2 |
|  | Inconclusive | 5 | 8 | 1 | 14 |
|  | Missing | 1 | 0 | 0 | 1 |
|  | Total | 70 | 176 | 14 | 260 |

## Acronyms

CCS: Clinician-Collected Swabs
CI: Confidence Interval
DDT: Daye Diagnostic Tampon
HPV: Human Papillomavirus
PPV: Positive Predictive Value
NPV: Negative Predictive Value
VSS: Vaginal Self-Swabs

